# The effect of migration, location in mining or borderland areas on HIV incidence among people who use drugs attending a harm reduction programme in Myanmar, 2014-2021: a retrospective cohort study

**DOI:** 10.1101/2023.12.05.23299510

**Authors:** Lucy Platt, Khine Wut Yee Kyaw, Sujit D. Rathod, Aung Yu Naing, Murdo Bijl, Bayard Roberts

## Abstract

**Background:** High HIV prevalence has been documented among people who use drugs (PWUD) in Myanmar particularly in mining and borderland areas. We estimated incidence of HIV among PWUD (via injecting and other routes) and examine associations between location in mining or borderland areas, migration and risk of infection.

**Methods and findings:** Analysis of data among PWUD registered at harm reduction programmes across Sagaing region, Kachin, and Northern Shan States between 2014-2021. Data on sociodemographic, drug use characteristics and clinic-level data on borderland or mining locations were collected at time of registration. Characteristics, repeat HIV testing and HIV seroconversion were analysed using a cohort approach and Poisson regression models examining associations between location in a borderland or mining area, migration and incidence of HIV, adjusting for confounders. Data were available from 85093 clients, 52526 reported HIV tests and 20.0% were seropositive. 38670 clients had no or only one recorded HIV result. The median time between HIV tests was 1.1 years. Among 13,359 clients with 2 or more HIV tests the HIV seroconversion rate was 3.8 per 100 person years (pyrs) (95% CI 3.6-4.0). Incidence among those who injected drugs was 6.9 per 100/pyrs, 8.9 among those aged ≤ 25 years, 2.3 among women, 2.3 among those who had migrated, 5.6 among those located in border areas, and 3.7 among those in mining areas. After adjusting for confounders HIV incidence remained higher for people located in borderland areas (Incidence Rate Ratio 1.67 95% CI 1.13-2.45) and lower among those who had migrated (IRR 0.56, 95% CI (0.39-0.82). There was no evidence of association between location in a mining area and HIV seroconversion.

**Conclusions:** Findings highlight the need to intensify harm reduction interventions with a focus on cross-border interventions. Increasing uptake of HIV testing alongside the scale up of evidenced based interventions to address sexual and injecting risk practices including PrEP, distribution of condoms, needles/syringe distribution and opioid agonist therapies is urgently needed to curb the high rates of HIV transmission among PWUD particularly among young people.

**Author summary:** *Why was this study done?:*

- Increased availability of drugs in producer countries or along trafficking routes alongside political instability and reduced enforcement has been linked to elevated drug use and outbreaks of HIV infection.
- Few studies focus on the extent to which structural factors, that is, political, social, or physical aspects of the environment, are associated with HIV infection in Myanmar though elevated HIV prevalence has been observed in rural and borderland areas.
- This study contributes to the limited evidence base on HIV incidence in South East Asia and Myanmar specifically.

*What did the researchers do and find?:*

- This study measures HIV incidence among people who use drugs using routine programmatic data and provides estimates of differential HIV risk associated with location in borderland and mining areas and experience of migration.
- We estimate HIV incidence to be 3.8 per 100 person years among people who use drugs, with higher incidence among those who inject and younger ages (<25 years)
- We identify people who use drugs registered at services in borderland areas to be at elevated risk of HIV acquisition and migrants at decreased risk.

*What do the findings mean?:*

- Findings point to the imperative for expanding HIV prevention interventions among PWUD, with a focus on cross-border interventions and addressing injecting and sexual risk practices.
- Findings support the emerging body of evidence highlighting the utility of programmatic data to estimate HIV incidence among key populations.
- Understanding differential risk in infections among people who use drugs and structural determinants is key to creating enabling environments through which effective HIV treatment and prevention interventions can be delivered.

## Introduction

Approximately 275 million people use drugs globally, between 11 and 21 million inject, and 20 million use amphetamine type stimulants (ATS).[1, 2] An estimated 4 million people who inject drugs (PWID) live in East and South-East Asia, representing a substantial proportion of the global population.[3] Use of ATS is increasing with use and production prominent in South East Asia.[4] Myanmar is one of the largest producers of ATS in the South East Asian region and the world’s second largest producer of opium.[5] Increased availability of drugs in producer countries or along trafficking routes alongside political instability and reduced enforcement has been linked to elevated drug use and outbreaks of HIV infection.[6, 7] Within Myanmar, drug production areas such as Kachin State, Northern Shan State, and the Sagaing region have the highest prevalence of drug use among the population.[1]

Between 1.4-2.8 million PWID are living with HIV globally.[2, 8] In 2020, 9% of all new HIV infections were among PWID and increasing to 20% if excluding African countries.[8] HIV incidence among PWID is estimated to be between 2.5-3 per 100 person years (pyrs) in Europe and North America and declining. However in many parts of South East Asia, Russia and Eastern Europe incidence is increasing, contributing between 30 and 39% of all new HIV infections in these regions in 2021.[3, 8, 9] A recent retrospective cohort study estimated incidence among PWID in Kachin State in Myanmar to be 7.1 per 100/pyrs, declining from 19.1 in 2008-2011 to 5.2 in 2017-2020.[10] Increased burden of HIV associated with injecting drugs compared to other routes of administration (e.g. insufflation, smoking) has focused HIV research and programmes on injecting practices. The prevalence of HIV associated with non-injecting drug use varies, depending on multiple factors including levels of unprotected sex within the population, engagement in sex work, the extent of sexual networks across injecting and other drug using communities as well as rates of transition between injecting and non-injecting practices.[11] In Vietnam HIV prevalence was 6.3% among young people (15-24 years) predominantly using methamphetamines and higher (15%) among those with a history of injecting.[12] In Myanmar, people using methamphetamines in Shan State frequently reported inconsistent condom use and multiple sex partners pointing to the potential for sexual transmission of HIV. [7]

There is a growing body of evidence documenting high HIV prevalence (34.9%) among PWID in Myanmar ranging from 7.6% to 61% and higher in rural areas of Bhamo and Waingmaw (61-56%) in Kachin State.[13] Elevated HIV prevalence in rural and borderland areas has been attributed to increased availability of drugs alongside higher rates of injecting across borders, equipment sharing practices and reduced access to harm reduction services.[13-15] Migration to and from rural and borderland areas is common with people migrating for work in agriculture, mining, sex work, or as a result of forced displacement.[7] Some evidence suggests higher risk sexual practices including condomless sex, multiple partners or history of sexually transmitted infections, are more common among migrating women using drugs in Muse, Myanmar on the border with China.[16] The ‘risk environment’ concept, developed to understand drug-related harms examines different types (physical, social, economic and political) and levels of influence (e.g. individual, community or national) in line with broader efforts to address structural determinants of health.[17, 18] This concept has been used to show how laws, housing, economic insecurity, stigma, displacement interplays with community factors (e.g. policing practices, access to services), to shape individual practices (e.g. sharing needles/syringes) and increase vulnerability to HIV among PWID.[11]

There has been little consideration of structural factors and HIV prevalence or incidence in Myanmar, although understanding incidence and differential risk is essential for targeting timely prevention interventions to populations and areas in need, as well as understanding impact of programmes over time. We undertook an analysis of programme data from a harm reduction service Myanmar with the aim to estimate the incidence of HIV among people who use drugs (PWUD) (both through injecting and other routes) and examine how physical aspects of the risk environment in the form of location in mining or borderland areas as well as migration might affect risk of infection.

## Methods

### Setting

Myanmar has suffered protracted armed conflict leading to large-scale forced displacement with millions moving within Myanmar as internally displaced persons or into neighboring countries as refugees. A total of 912,000 people are internally displaced across Myanmar, with the largest populations in Kachin, Chin, Shan and Rakhine.[19] Sagaing region, Kachin and Northern state have vast borderland areas adjoining India, China, Laos and Thailand, primarily composed of marginalized ethnic groups. Mountains and dense forest terrain limit accessibility to health services, high prevalence of HIV and outbreaks of malaria have been documented.[20, 21] Evidence shows elevated opium, heroin or ATS use among those working in mining industries in Myanmar as a strategy to cope with difficult working conditions.[7] Borderland and mining areas are characterized by high levels of sex work and population mobility.

The Asian Harm Reduction Network (AHRN) has been providing harm reduction services in Kachin state since 2003 and expanded to Northern Shan State and Sagaing region under the National AIDS Programme guidelines for the treatment and prevention of HIV among key populations. Attendance is voluntary and services include provision of needles and syringes; condom and lubricant distribution; opioid agonist therapy (OAT); HIV testing and counselling; antiretroviral therapy (ART) provision, information, education and communication; Hepatitis C and B testing and treatment; Hepatitis B vaccination; sexually transmitted infection (STI) and Tuberculosis (TB) prevention, diagnosis and treatment; and mental health assessment, treatment and referral. Aluminium foil is distributed for smoking of opium or heroin to encourage transition away from injecting or to reduce sharing smoking equipment. Clients consist of people who use or inject drugs, their sexual partners and family members. For this analysis we focus on people who currently use drugs. We focus on individuals: i) with one or more HIV test results; ii) who tested HIV negative at first HIV test; and iii) who registered since 2014 when ART scale-up was intensive and routine HIV testing was increased.

### Study design and data collection

We conducted longitudinal analysis of routine data collected from 35 of AHRN’s project sites across 22 townships in Myanmar between January 2014 and December 2021. Deidentified data were extracted for analysis in January 2022. Project sites include fixed sites (drop-in centres) and through mobile teams to reach the largely rural populations. New clients are provided with a unique identifier and complete a standardized registration form. This is completed on paper at any service (mobiles, via outreach, at drop-in centres (DIC)) and then entered into an electronic database. Questions on drug use (type and mode) and other demographic characteristics are recorded during registration.

HIV testing is provided at fixed site DICs and through mobile medical teams with a suggested frequency of six months. HIV testing is encouraged through outreach workers and via peer workers. Counsellors (nurses or peers) provide pre-test counselling, and after obtaining consent, whole blood specimens (finger prick or venipuncture) are taken in DICs or mobile clinics and tested using Alere Determine HIV 1-2 (Alere Medical Company Ltd, Japan). Reactive samples are retested at laboratories (for DICs) or at DICs (following community testing) using confirmatory tests conducted in parallel Unigold (Trinity Biotech Manufacturing Ltd., Ireland) and Stat-pak (Chembio Diagnostic Systems Ltd., USA) with post-test counselling provided at return of results. Clients with confirmed HIV positive results are referred to AHRN ART satellite sites for pre-ART assessment, co-trimoxazole preventive therapy, opportunistic infections screening, treatment and counselling.

### Covariables

Key exposures were indicators of geographical context including: (i) migration; (ii) location of clinics in borderland; or (iii) mining areas. Location in borderland or mining areas were extracted from profiles of townships, crossed checked with project staff and attributed to individuals according to their location at registration (at DIC or outreach).[22] Experience of migration was self-reported defined as living away from hometown and movement to various locations for three months or more. Clinics were classified as being in the states of Kachin, or Shan (North) or Sagaing Region.

We considered other factors associated with HIV risk including gender (male/female); education (illiterate, primary/literate, completed secondary and tertiary or more); age (≤25, 25-34, 35-44, ≥45 years); marital status (single, married, widowed/divorced); history of injecting (yes/no). Drug use (current) was grouped into heroin (yes/no), amphetamines, other drugs (including alcohol and methadone maintenance therapy), with clients being able to report multiple drugs. Occupation was recoded from an open-text response into nine categories. We defined a missing data category for covariates with >10% missing data. All indicators represent characteristics reported at the time of client registration.

### Outcome

HIV seroconversion was defined as a positive test result on a date after a negative result, per AHRN clinical records. We included clients who had an HIV negative test result followed by one or more completed test. Applying methods for estimating HIV incidence using routine clinic records[23], we calculated HIV-negative survival time for each client, starting with the registration date, when exposures and covariable data were collected. Survival time for clients who remained seronegative ended on the date of their last HIV negative test result and for those that seroconverted at the midpoint of their last HIV negative result date and their first HIV positive result date.

### Data analysis

We describe the client population of people who use drugs served by AHRN between January 2014 and September 2021 stratified by exclusion criteria (no HIV test; testing HIV positive at first test; and only 1 HIV test) in order to assess potential selection bias of the sample included in the HIV incidence analysis. We use medians and IQRs for continuous variables and counts and percentages for categorical variables.

We present the sample included in the HIV incidence analysis stratified by our three exposures of interest (migration, location in borderland or mining area). We estimated crude incidence rate ratios for our exposures and other covariables on HIV incidence using separate univariable Poisson regression models. Finally, we estimated incidence rate ratios for the exposures, using separate multivariable Poisson regression models with adjustment for potential confounders. All models adjusted for age, education, and registration year. For the effect of being in a border area, we adjusted for gender, state of clinic, and mining catchment area and for being in a mining catchment area, we adjusted for gender additionally. Confounders were selected based on the available covariables in the dataset which were imbalanced by exposure status, were potential risk factors for HIV, and unlikely to be on the causal pathway between the exposure and outcome. We did not adjust for marital status, migration, or occupation due to the high level of missing data for these measures. Poisson regression models included the log survival time as an offset, and had standard errors adjusted for project sites (mobile and DIC) as clusters. Participants with missing exposure data were excluded from analyses involving that exposure. We used Stata 18 (Stata Corp) in all analyses.

### Ethics

Ethical approval was obtained from LSHTM research ethics committee (Ref: 22838).

## Results

Between January 2014 and September 2021, there were 85093 people registered across 35 AHRN project sites and 22 townships (Table 1). The median age was 38 years, 95.6% were male, 4.6% had not completed any school/were illiterate, and 18.4% were married. While 15.8% identified as a migrant, another 41.5% had migration status missing. Occupation was diverse, 32.0% worked in agriculture, 24.3% in mining industries and 6.0% were students or had no income, 25.3% of occupation status missing. Heroin was used by 93.9% of clients, 59.8% used amphetamines, and 48.9% used opium. A total of 47,538 (59.5%) used both heroin and ATS (data not shown). Overall 48.5% injected drugs. For service registration location, 19.5% were registered in a border area while 76.2% were registered in a mining area. We excluded 71734 clients from the HIV incidence analysis for the following reasons: no HIV test (n=32567, 38.3%), lack of a follow-up HIV test (n=27040, 32.8%), initial HIV positive result (n=11630, 13.7%), and due to inability to match names to HIV results or discrepancies between test date and registration data (n=497, 0.5%) (Figure 1).

**Figure 1:**
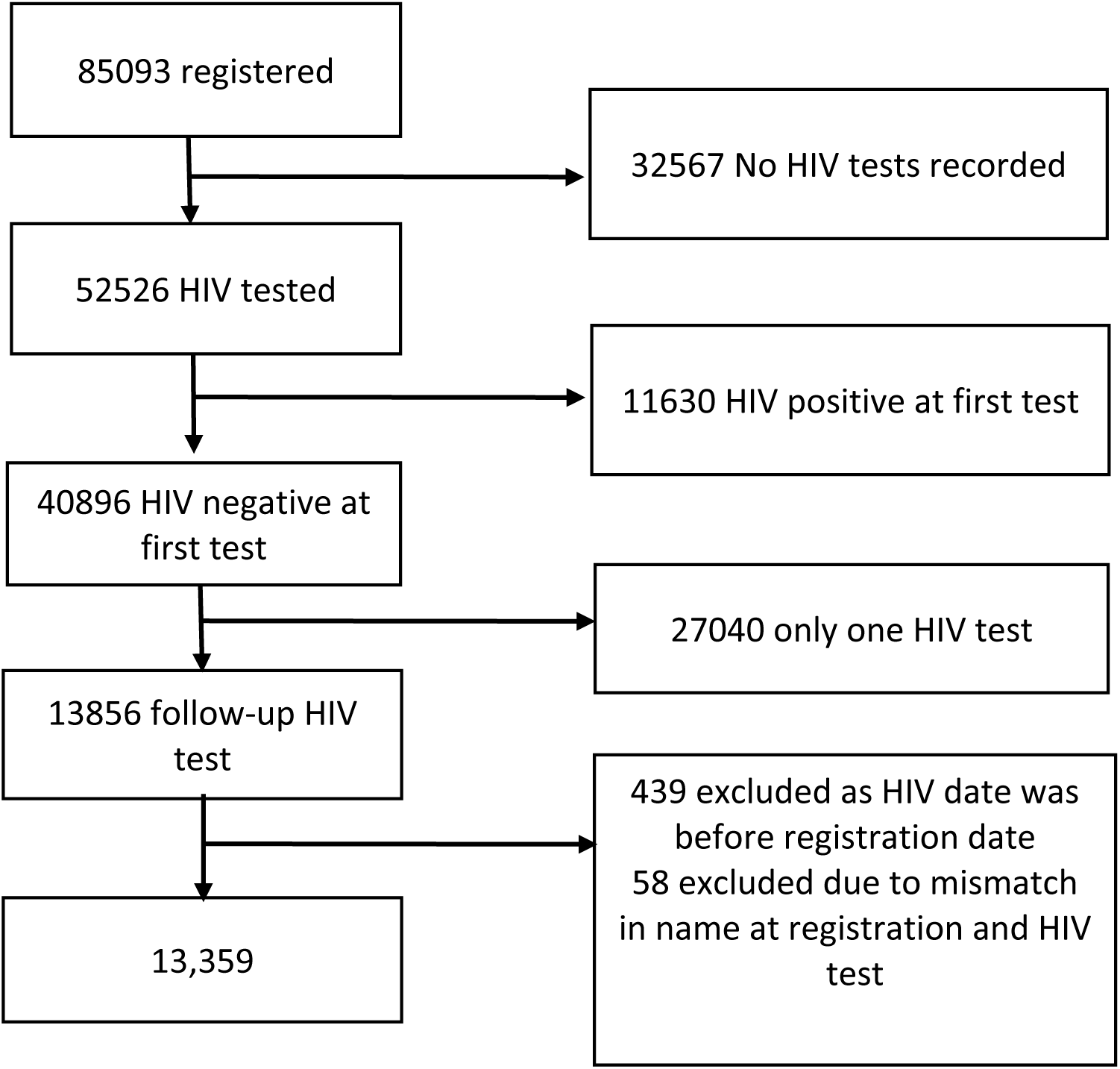
HIV incidence analysis exclusions among AHRN clients in Myanmar, 2014-2021.

**Table 1.** Characteristics of newly registered AHRN clients by HIV incidence analysis inclusion status, Myanmar, 2014-2022.

|  | Total | Excluded participants* |  |  | Included |
| --- | --- | --- | --- | --- | --- |
| Variable |  | No HIV test | HIV+ at first test | 1 HIV test (HIV-) |  |
|  | n(%) row | n(%) row | n(%) row | n(%) row | n(%) row |
| **Total** | 85093 (100) | 32567 (38.3) | 11630 (13.7) | 27040 (32.8) | 13359 (15.7%) |
| **Age, median (IQR)** | 38 (31-46) | 36 (30-43) | 33 (28-40) | 36 (29-44) | 36 (30-43) |
|  | n(%) col | n(%)col | n(%)col | n(%)col | n(%)col |
| <=25 | 6683(7.8) | 2091 (6.4) | 944 (8.1) | 2709 (10.0) | 693 (5.2) |
| 25-34 | 32550 (38.2) | 12565 (38.6) | 5647 (48.6) | 9887 (36.6) | 4287 (32.0) |
| 35-44 | 27256(32.0) | 10782 (33.1) | 3594 (30.9) | 8077 (29.9) | 4648 (34.8) |
| >=45 | 18648 (21.9) | 6989 (21.5) | 1444 (12.4) | 6340 (23.4) | 3730 (27.9) |
| **Gender** |  |  |  |  |  |
| Male | 81314 (95.6) | 31229 (95.9) | 11323 (97.4) | 25553 (94.5) | 12736 (95.3) |
| Female | 3779 (4.4) | 1338 (4.1) | 307 (2.6) | 1487 (5.5) | 623 (4.6) |
| **Education** |  |  |  |  |  |
| Illiterate/other | 3909(4.6) | 1386 (4.3) | 501 (4.3) | 1517 (5.6) | 498 (3.7) |
| Primary/literate | 24258 (28.5) | 8051 (24.7) | 3406 (28.3) | 8102 (30.0) | 4511 (33.8) |
| Up to secondary | 43660 (51.3) | 15124 (46.4) | 6764 (58.2) | 14345 (53.0) | 7173 (53.7) |
| Tertiary or more | 3283 (3.8) | 1253 (3.8) | 311 (2.7) | 1141 (4.2) | 556 (4.2) |
| Missing | 9983 (11.7) | 6753 (20.7) | 648 (5.6) | 1935 (7.2) | 621 (4.6) |
| **Marital** |  |  |  |  |  |
| Post-marital | 3024 (3.5) | 576 (1.8) | 614 (5.3) | 1249 (4.6) | 555 (4.1) |
| Married | 15684(18.4) | 3078 (7.4) | 1738 (14.9) | 7431 (27.5) | 3293 (24.6) |
| Single | 11271 (13.2) | 2221 (6.8) | 2118 (18.2) | 5060 (18.7) | 1780 (13.3) |
| Missing | 55114(64.7) | 26692 (82.0) | 7160 (61.6) | 13300 (49.2) | 7731 57.8) |
| **Occupation** |  |  |  |  |  |
| Agriculture | 27240 (32.0) | 8819 (27.1) | 4078 (35.1) | 9486 (35.1) | 4631(34.7) |
| Driver | 3291 (3.9) | 1421 (4.4) | 324 (2.8) | 985 (3.6) | 536 (4.0) |
| Drug/casino/sex | 1239 (1.5) | 406 (1.2) | 103 (0.9) | 446 (1.6) | 270 (2.0) |
| Mining | 20642 (24.3) | 6616 (20.3) | 3267 (28.1) | 6874 (25.4) | 3798 (28.4) |
| Student / no income | 5110 (6.0) | 1923 (5.9) | 828 (7.1) | 1604 (5.9) | 735 (5.5) |
| Casual work | 3758 (4.4) | 1547 (4.7) | 499 (4.3) | 1081 (4.0) | 602 (4.5) |
| Uniformed officer | 17 (0.0) | 6 (0.2) | 3 (0.03) | 8 (0.03) | 0 (0.0) |
| Skilled / office | 2240(2.6) | 830 (2.5) | 279 (2.4) | 795 (2.9) | 326 (2.4) |
| Other / missing | 21556 (25.3) | 10999 (33.8) | 2249 (19.3) | 5761 (21.3) | 2461 (18.4) |
| **Moved away from home** |  |  |  |  |  |
| No | 36311(42.7) | 13793 (42.3) | 4569 (38.3) | 12257 (45.7) | 5319 (39.8) |
| Yes | 13426 (15.8) | 4229 (13.0) | 1559 (13.4) | 5541 (20.5) | 2043 (15.3) |
| Missing | 35356 (41.5) | 14546 (44.7) | 5502 (47.3) | 9142 (33.8) | 5997 (44.9) |
| **Drug use** |  |  |  |  |  |
| Heroin (yes vs no) | 79930 (93.9) | 30547 (93.8) | 11437 (98.3) | 24569 (90.9) | 12897 (96.5) |
| Opium (yes vs no)s | 41600 (48.9) | 14375 (44.1) | 6180 (53.1) | 13408 (49.6) | 7360 (55.1) |
| Amphetamines (yes vs no) | 50887 (59.8) | 18069 (55.5) | 6700 (57.6) | 17158 (63.4) | 8645 (64.7) |
| Other drug<sup>s</sup>(yes vs no) | 6579 (7.7) | 2212 (6.8) | 1057 (9.1) | 2006 (7.4) | 1248 (9.3) |
| Injects drugs (yes vs no) | 41309 (48.5) | 14734 (45.2) | 10322 (88.7) | 9860 (36.5) | 6129 (45.8) |
| **Type of clinic** |  |  |  |  |  |
| Fixed site drop-in centre | 74352 (87.4) | 29564 (90.8) | 9957 (85.6) | 23079 (85.3) | 11333 (84.8) |
| Mobile | 10741 (12.6) | 3003 (9.2) | 1673 (14.4) | 3961 (14.6) | 2026 (15.2) |
| **Clinic in border area** |  |  |  |  |  |
| No | 67854(79.7) | 26366 (81.0) | 8504 (73.1) | 20928 (77.4) | 11608 (86.8) |
| Yes | 16627 (19.5) | 5706 (17.5) | 2083 (26.5) | 6038 (22.3) | 1751 (13.1) |
| **Clinic in mining area** |  |  |  |  |  |
| No | 19535 (23.0) | 9132 (28.0) | 1722 (14.8) | 5597 (20.7) | 2909 (21.8) |
| Yes | 64863 (76.2) | 22929 (70.4) | 9864 (84.8) | 21310 (78.8) | 10438 (78.1) |
| **Clinic State/Region** |  |  |  |  |  |
| Kachin | 49925 (59.6) | 18892 (58.0) | 7907 (68.0) | 15730 (58.2) | 7183 (53.8) |
| Sagaing | 27732 (32.6) | 10456 (32.1) | 3200 (27.5) | 8741 (32.3) | 5071 (38.0) |
| Shan (North) | 6824 (8.0) | 2724 (8.4) | 480 (4.1) | 2495 (9.2) | 1105 (8.3) |
| **HIV test** |  |  |  |  |  |
| Negative | 40896 (48.1) | 0 (0.0) | 0 (0.0) | 27040 (100) | 13259 (100) |
| Positive | 11630 (16.2) | 0 (0.0) | 11630 (100) | 0 (0.0) | 0 (0.0) |
| Missing | 32567(38.3) | -32567 (100) | 0 (0.0) | 0 (0.0) | 0 (0.0) |
| **Date of registration** |  |  |  |  |  |
| 2014 | 5275 (6.2) | 2969 (9.1) | 687 (5.9) | 807 (3.0) | 779 (5.8) |
| 2015 | 5275 (6.2) | 2670 (8.2) | 759 (6.5) | 896 (3.3) | 929 (6.9) |
| 2016 | 7978 (9.4) | 3859 (11.8) | 953 (8.2) | 1565 (5.8) | 1571 (11.7) |
| 2017 | 9229 (10.8) | 4542 (14.0) | 1202 (10.3) | 1899 (7.0) | 1565 (11.7) |
| 2018 | 12121 (14.4) | 4550 (14.0) | 1941 (16.7) | 3310 (12.2) | 2268 (17.0) |
| 2019 | 17014 (20.0) | 5487 (13.7) | 2651 (22.8) | 5453 (20.2) | 3312 (24.8) |
| 2020 | 16098 (18.9) | 4451 (13.7) | 2160 (18.6) | 6701 (24.8) | 2642 (19.8) |
| 2021 | 12103 (14.2) | 4039 (12.4) | 1278 (11.0) | 6409 (23.7) | 293 (2.2) |

\*497 (0.5%) people were excluded due to mismatch in names at registration and HIV test or if HIV test date was prior to registration date. All indicators were collected at point of client registration <sup>s</sup> Other drugs included, marijuana, formula, diazepam, alcohol and methadone.

The median age of included participants was younger than the total sample (36 vs 38 years) and those people who had no HIV test were marginally younger (36 years). Proportionally more participants reporting migration had only one HIV test compared to the total sample included (20.% vs15.8%). There were higher levels of opium use among included participants compared to the total sample (55.1% vs 48.9%) and amphetamine use (64.7% vs 59.8%). Proportionally fewer participants were registered in borderland areas in the final analytical sample (13.1%) compared to the total sample (19.5%). A higher proportion were excluded due to HIV positive first test (26.%) and no follow-up HIV test (22.3%).

### Characteristics by migration, registration in borderland or mining area

Figure 2 depicts the geographical distribution of townships in which AHRN services operate according to location in borderland or mining areas and inclusion in the analysis. We included data from 15/22 townships, of which 8 were located in mining areas, 3 were in borderland areas, 2 were in both mining and borderland areas and 2 were in neither a borderland nor mining area. Townships were excluded where projects sites did not work with people who use drugs and one site conducted HIV testing only but did not record any data.

**Figure 2:**
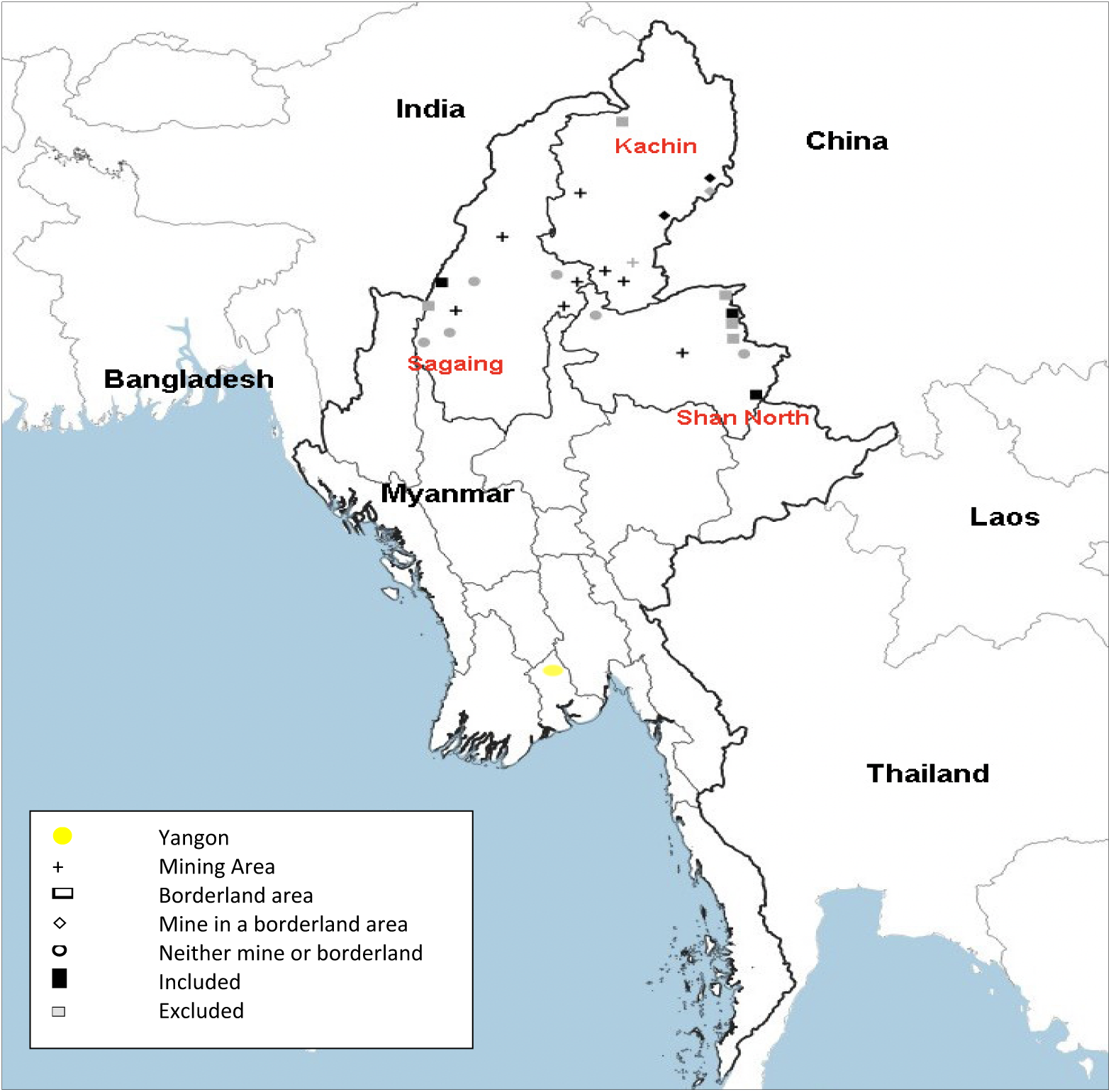
Distribution of townships included in the analysis in which AHRN provide harm reduction services (mobile or fixed sites) according to their location in borderland or mining areas.

Figure 1: Distribution of townships included in the analysis in which AHRN provide harm reduction services (mobile or fixed sites) according to their location in borderland or mining areas

Table 2 summarises the characteristics of clients by migrant status, registration in border area vs non-border area and registration in a mining area vs non-mining area. Clients who identify as migrants were more likely to be single (33.0%) than non-migrants (18.6%). Migrants were more likely to use amphetamines (73.4% vs 55.3%), and to register at an AHRN clinic in a mining area (96.4% vs 73.0%) compared to non-migrants. Clients who registered at an AHRN clinic in border areas were more likely to be married (37.5%) than clients who registered elsewhere (22.9%), to be illiterate (8.3% vs 3.0%), to work in agricultural labour (52.4% vs 31.9%) to inject drugs (54.4% vs 44.4%), and were less likely to be a migrant (56.3% vs 81.5%) or use amphetamines (54.8% vs 66.0%). Clients who registered at an ARHN clinic in a mining area were more likely to be female (5.6%) than clients who registered elsewhere (2.2%), less likely to use opium (51.0% vs 69.7%) and were more likely to be a migrant (18.9% vs 2.1%)

**Table 2:** Characteristics of included clients by migration experience, location in borderland or mining areas (n=13359)

| Variable | Migrant |  | Borderland area |  | Mining area |  |
| --- | --- | --- | --- | --- | --- | --- |
|  | No | Yes | No | Yes | No | Yes |
| Total* | 5319 | 2043 | 11632 | 1772 | 2919 | 10473 |
| **Demographic characteristics** |  |  |  |  |  |  |
| Age, median (IQR) | 38 (31-47) | 37 (31-44) | 38 (32-46) | 36 (30-44) | 38 (31-46) | 38 (31-46) |
| **Gender** |  |  |  |  |  |  |
| Male | 5127 (95.9) | 1914 (93.2) | 11079 (95.3) | 1662 (93.8) | 2856 (97.8) | 9884 (94.4) |
| Female | 219 (4.1) | 140 (6.8) | 553 (4.8) | 110 (6.2) | 63 (2.2) | 589 (5.6) |
| **Education** |  |  |  |  |  |  |
| Illiterate/other | 144 (2.7) | 63 (3.1) | 348 (3.0) | 147 (8.3) | 77 (2.6) | 417 (4.0) |
| Primary/literate | 2160 (40.4) | 555 (27.0) | 4088 (35.1) | 443 (25.0) | 1122 (38.4) | 3407 (32.5) |
| Up to secondary | 2557 (47.8) | 1161 (56.5) | 6166 (53.0) | 1037 (58.5) | 1519 (52.0) | 5676 (54.2) |
| Tertiary or more | 180 (3.4) | 71 (3.5) | 478 (4.1) | 76 (4.3) | 147 (5.0) | 406 (3.9) |
| Missing | 305 (5.7) | 204 (9.9) | 552 (4.8) | 69 (3.9) | 54 (1.8) | 567 (5.4) |
| **Marital** |  |  |  |  |  |  |
| Post-marital | 273 (5.1) | 265 (12.9) | 484 (4.2) | 73 (4.1) | 86 (2.9) | 467 (4.5) |
| Married | 2405 (45.0) | 697 (33.9) | 2665 (22.9) | 664 (37.5) | 670 (22.9) | 2657 (25.4) |
| Single | 992 (18.6) | 677 (33.0) | 1495 (12.8) | 289 (16.3) | 258 (8.8) | 1520 (14.5) |
| Missing | 1676 (31.3) | 415 (20.2) | 6988 (60.1) | 746 (42.1) | 1905 (65.3) | 5829 (55.7) |
| **Occupation** |  |  |  |  |  |  |
| Agriculture | 2836 (53.2) | 139 (6.7) | 3716 (31.9) | 928 (52.4) | 1934 (66.3) | 2710 (25.9) |
| Driver | 197 (3.7) | 71 (3.4) | 484 (4.2) | 51 (2.9) | 98 (3.4) | 437 (4.2) |
| Drug/casino/sex | 61 (1.1) | 80 (3.9) | 233 (2.0) | 47 (2.6) | 45 (1.5) | 224 (2.1) |
| Mining | 505 (9.5) | 1213 (58.7) | 3773 (32.4) | 26 (1.5) | 47 (1.6) | 3752 (35.8) |
| Student/no income | 273 (5.1) | 80 (3.9) | 561 (4.8) | 190 (10.7) | 156 (5.4) | 595 (5.7) |
| Casual | 281 (5.3) | 77 (3.7) | 506 (4.3) | 100 (5.6) | 194 (6.6) | 412 (3.9) |
| Skilled / office | 152 (2.8) | 31 (1.5) | 238 (2.0) | 86 (4.9) | 128 (4.4) | 196 (1.9) |
| Other / missing | 1028 (19.3) | 374 (18.1) | 2118 (18.2) | 343 (19.4) | 315 (10.8) | 2145 (20.5) |
| **Migration/moved away from home** |  |  |  |  |  |  |
| No | - | - | 4573 (39.3) | 773 (43.6) | 1446 (49.5) | 3900 (37.2) |
| Yes | - | - | 1935 (16.6) | 119 (6.7) | 61 (2.1) | 1981 (18.9) |
| Missing | - | - | 5124 (44.0) | 880 (49.7) | 1412 (48.4) | 4592 (43.9) |
| **Drug use** |  |  |  |  |  |  |
| Heroin (yes vs no) | 5196 (97.2) | 1924 (93.7) | 11241 (96.6) | 1650 (93.1) | 2850 (97.6) | 10041 (95.9) |
| Amphetamine (yes vs no) | 2958 (55.3) | 1507 (73.4) | 7672 (66.0) | 972 (54.8) | 2108 (72.2) | 6524 (62.3) |
| Opium (yes vs no) | 3012 (56.4) | 764 (37.1) | 6502 (55.9) | 873 (49.3) | 2038 (69.9) | 5337 (50.9) |
| Other (yes vs no) | 955 (17.9) | 61 (3.0) | 1248 (10.7) | 82 (4.6) | 116 (4.0) | 1214 (11.6) |
| Injects drugs(yes vs no) | 2344 (43.9) | 892 (43.4) | 5168 (44.4) | 964 (54.4) | 1266 (43.4) | 4866 (46.5) |
| **Clinic location** |  |  |  |  |  |  |
| **State/Region** |  |  |  |  |  |  |
| Kachin | 1431 (43.7) | 1843 (56.3) | 6214 (53.4) | 999 (56.4) | 0 (0.0) | 7213 (68.9) |
| Sagaing | 3633 (97.6) | 91 (2.4) | 4541 (39.0) | 540 (30.5) | 2699 (92.4) | 2383 (22.7) |
| Shan (North) | 270 (67.5) | 130 (32.5) | 876 (7.5) | 233 (13.1) | 221 (7.6) | 876 (8.4) |
| **In border area** |  |  |  |  |  |  |
| No | 4573 (85.5) | 1935 (94.2) |  |  | 2157 (73.9) | 9475 (90.5) |
| Yes | 773 (14.5) | 119 (5.8) |  |  | 762 (26.1) | 998 (9.5) |
| **In mining area** |  |  |  |  |  |  |
| No | 1446 (27.0) | 61 (3.0) | 2157 (18.5) | 762 (43.0) |  |  |
| Yes | 3900 (73.0) | 1981 (96.4) | 9475 (81.5) | 998 (56.3) |  |  |
| Missing | 0 (0.0) | 12 (0.6) | 0 (0.0) | 12 (0.7) |  |  |

\*Missing data: Migrants (n=6007, 44.8%), Borderland (n=1771,13.2%), Mining (n=12 0.1%)

### HIV incidence rate and associations with demographic characteristics

Of the 13,359 included clients, there were 29,491 person-years of follow up (median 1.7 years, IQR 0.9-3.1) recorded between 2014 and 2021. A total of 33,022 HIV tests were conducted (median 2 IQR 2-3 tests per participant and 1.1 years between test); 1,114 clients had an HIV-positive follow-up test and another 12,245 had only HIV-negative follow up results, corresponding to an HIV incidence rate of 3.8/100 person-years. Overall, between 2014-2021 there were 12,736 men who registered at AHRN clinics, contributing 28,251 person-years of observation, 1,085 HIV seroconversions, and an HIV incidence rate of 3.8 per 100 person-years, compared to 2.3 for women.

The incidence rate ratio (IRR) for women compared to men was 0.61 (95% CI of 0.38-0.97). We also observed lower HIV incidence among clients who were older compared to those <=25 years (IRR >=45 years 0.17 95% CI 0.12-0.25; 35-44 years= 0.40 95% CI 0.28-0.57; 25-34 years=0.65 95% CI 0.48-0.87), migrants compared to non-migrants (IRR 0.55 95% CI 0.37-0.82) or married compared to widowed/divorced (IRR 0.63 95% CI 0.46-0.83). We observed greater HIV incidence among clients with higher educational attainment, who used heroin compared to those who did not (3.11 95% CI 2.27-4.27), used drugs via injection versus non-injecting (IRR 5.5 95% CI 4.25-7.04), or were registered at a clinic in a border area versus not (IRR 1.6 95% CI 0.97-2.62). Results are summarised in Table 3.

**Table 3:** Client characteristics and crude association with HIV incidence rate among AHRN clients who use drugs, 2014-2022 (n=13,359)

| Variable | incident HIV | Total at start of observation | Person-years of survival | Rate/100 pyrs* | Incidence Rate Ratio (95%CI) <sup>a</sup> |
| --- | --- | --- | --- | --- | --- |
| **Total** | 1114 | 13359 | 29491 | 3.8 |  |
| **Age** |  |  |  |  |  |
| $\leq 25$ | 87 | 693 | 981 | 8.9 | 1.0 |
| 25-34 | 502 | 4307 | 8698 | 5.3 | 0.65 (0.48, 0.87) |
| 35-44 | 390 | 4655 | 10964 | 3.2 | 0.40 (0.28, 0.57) |
| $\geq 45$ | 135 | 3740 | 8847 | 1.3 | 0.17 (0.12, 0.25) |
| **Gender** |  |  |  |  |  |
| Male | 1085 | 12736 | 28251 | 3.8 | 1.0 |
| Female | 29 | 623 | 1240 | 2.3 | 0.61 (0.38, 0.97) |
| **Education** |  |  |  |  |  |
| Illiterate/other | 30 | 498 | 1140 | 2.6 | 1.0 |
| Primary/literate | 350 | 4511 | 9432 | 3.7 | 1.39 (1.06, 1.89) |
| Up to secondary | 679 | 7173 | 16651 | 4.0 | 1.50 (1.19, 1.96) |
| Tertiary or more | 44 | 556 | 1370 | 3.2 | 1.21 (0.83, 1.80) |
| Missing | 28 | 621 | 898 | 2.2 | 0.83 (0.40, 1.76) |
| **Occupation** |  |  |  |  |  |
| Agriculture | 376 | 4631 | 8665 | 4.3 | 1.0 |
| Driver | 52 | 536 | 1438 | 3.6 | 0.83 (0.65, 1.06) |
| Drug/casino/sex | 18 | 270 | 611 | 2.9 | 0.68 (0.37, 1.22) |
| Mining | 310 | 3798 | 9253 | 3.3 | 0.77 (0.51, 1.17) |
| Students / no income | 77 | 735 | 1779 | 4.3 | 1.00 (0.77, 1.29) |
| Casual | 53 | 602 | 1375 | 3.8 | 0.89 (0.67, 1.18) |
| Skilled / office | 22 | 326 | 719 | 3.0 | 0.9 (0.46, 1.05) |
| Other / missing | 206 | 2461 | 5651 | 3.6 | 0.84 (0.66, 1.06) |
| **Marital status** |  |  |  |  |  |
| Post-marital | 53 | 555 | 993 | 5.3 | 1.0 |
| Married | 191 | 3293 | 5705 | 3.3 | 0.63 (0.46, 0.86) |
| Single | 147 | 1780 | 3027 | 4.8 | 0.91 (0.73, 1.13) |
| Missing | 723 | 7731 | 19765 | 3.6 | 0.68 (0.42, 1.11) |
| **Migration/moved away from home** |  |  |  |  |  |
| No | 388 | 5319 | 9196 | 4.2 | 1.0 |
| Yes | 94 | 2043 | 4014 | 2.3 | 0.55 (0.37, 0.82) |
| Missing | 632 | 5997 | 16280 | 3.9 | 0.92 (0.64, 1.33) |
| **Uses heroin** |  |  |  |  |  |
| No | 11 | 462 | 887 | 1.1 |  |
| Yes | 1103 | 12897 | 28604 | 3.8 | 3.11 (2.27, 4.27) |
| **Uses opium** |  |  |  |  |  |
| No | 461 | 5999 | 12673 | 3.6 | 1.0 |
| Yes | 653 | 7360 | 16818 | 3.9 | 1.07 (0.89, 1.28) |
| **Uses amphetamine** |  |  |  |  |  |
| No | 404 | 4714 | 10963 | 3.7 | 1.0 |
| Yes | 710 | 8645 | 18529 | 3.8 | 1.04 (0.86, 1.25) |
| **Uses injection drugs** |  |  |  |  |  |
| No | 198 | 7230 | 15976 | 1.2 | 1.0 |
| Yes | 916 | 6129 | 13516 | 6.8 | 5.5 (4.25, 7.04) |
| **Location of clinic** |  |  |  |  |  |
| Kachin | 618 | 7183 | 17598 | 3.5 | 1.0 |
| Sagaing | 421 | 5071 | 9348 | 4.5 | 1.28 (0.87, 1.88) |
| Shan (North) | 75 | 1105 | 1030 | 2.9 | 0.84 (0.56, 1.25) |
| **In border area** |  |  |  |  |  |
| No | 916 | 11608 | 25978 | 3.5 | 1.0 |
| Yes | 198 | 1751 | 3514 | 5.6 | 1.60 (0.97, 2.62) |
| **In mining area** |  |  |  |  |  |
| No | 259 | 2909 | 6427 | 4.0 | 1.0 |
| Yes | 855 | 10438 | 23054 | 3.7 | 0.92 (0.68, 1.24) |
| **Registration year** |  |  |  |  |  |
| 2014 | 125 | 779 | 3640 | 3.4 | 1.0 |
| 2015 | 123 | 929 | 3947 | 3.1 | 0.90 (0.67, 1.23) |
| 2016 | 147 | 1571 | 5593 | 2.6 | 0.76 (0.51, 1.14) |
| 2017 | 159 | 1565 | 4213 | 3.8 | 1.10 (0.74, 1.64) |
| 2018 | 227 | 2268 | 4571 | 5.0 | 1.44 (0.94, 2.21) |
| 2019 | 212 | 3312 | 4861 | 4.3 | 1.27 (0.75, 2.13) |
| 2020 | 116 | 2642 | 2492 | 4.7 | 1.35 (0.83, 2.20) |
| 2021 | 5 | 293 | 173 | 2.9 | 0.84 (0.29, 2.45) |

a Crude incidence rate ratio estimated with Poisson regression with log survival time as an offset and 95% CI adjusted for clustering by project site

Association between HIV incidence and migration, location in a borderland or mining area After adjusting for potential confounders, clients who were registered at a clinic in a border area had 67% higher incidence of HIV (IRR 1.67, 95% CI 1.13-2.45) relative to those who registered elsewhere. We did not observe a difference in the incidence of HIV for clients according to whether their clinic registration was in a mining area or not. Clients who identified as a migrant at registration had 44% lower incidence of HIV (IRR 0.56, 95%CI 0.39-0.82) relative to those who did not identify as a migrant (see Table 4).

**Table 4:** Geographic characteristics and adjusted association with HIV incidence among AHRN clients, 2014-2022.

| **Risk factor** | **Seropositive cases/Total** | **Adjusted Incidence Rate Ratio (95% CI) <sup>a</sup>** | **P value** |
| --- | --- | --- | --- |
| **Client in border area clinic** | 198/11608 | 1.67 (1.13, 2.45)<sup>b</sup> | 0.009 |
| **Client in mining area clinic** | 855/10438 | 0.93 (0.72, 1.20)<sup>c</sup> | 0.582 |
| **Client has migrated/moved away from home** | 94/2043 | 0.56 (0.39, 0.82)<sup>c</sup> | 0.003 |

<sup>a</sup> Incidence rate ratio estimated with Poisson regression with log survival time as an offset
<sup>b</sup> IRR adjusted for adjusted for age, gender, education, calendar year of registration, registration in mining area, state of clinic
<sup>c</sup> IRR adjusted for adjusted for age, gender, education, calendar year of registration
All 95% CIs adjusted for clustering by project site

## Discussion

Our study found high HIV incidence among people who use drugs (PWUD) accessing services through AHRN of 3.8 cases per 100 person-years and higher among those who inject (6.8/100 pyrs) and those aged 25 years or younger (8.9/100 pyrs). Higher seroconversion rates were observed among those registered in a border area (IRR 1.67 95% CI 1.13-2.45) and lower rates among migrants (IRR 0.56 95% CI 0.39-0.82) and there was no evidence of association with location in a mining area.

Findings support evidence of differential risk of HIV transmission among PWUD situated in border areas than adults in non-border areas.[21, 24-26] Research among people who inject heroin in borderland areas in remote parts of Shan State document frequent injection (at least daily) and high levels of sharing of needles/syringes.[7] Other evidence from Ruili immediately across the border in China, found that injecting drugs in both China and Myanmar was associated with increased odds of sharing injecting equipment and weaker evidence of an association with testing positive for HIV.[15] Elevated ATS use in borderland cities in proximity to ATS distribution has been observed both in the region and in border cities between Mexico and the United States.[24, 27] We found some differences in drug use between those located in borderland areas compared to non-borderland, ATS use was less frequently reported but a greater proportion of people injected drugs, had higher levels of illiteracy and worked in agriculture. This likely reflects the increased availability of heroin as well as poorer access to education and more limited employment opportunities in borderland areas. Structural barriers including the physical landscape (thick forests and mountains), historic armed conflict, insecure income and involuntary migration make it difficult accessing health services and other necessary infrastructures.[28] Proportionally fewer AHRN clients were registered at project sites in borderland areas (19.5%) compared to mining areas (76.2%). Assuming demand for services is comparable, this could indicate reduced access to harm reduction services that, alongside increased levels of injecting, may contribute to elevated incidence observed in borderland areas but not mining areas. Our findings suggest less ATS use and comparable prevalence of heroin or injecting drugs in mining areas compared to non-mining areas in contrast to reports that document intensive ATS and heroin use among people working in mines.[14] Given difficulties in providing services in sensitive areas such as borderlands and mines, further research is needed to better quantify existing coverage of harm reduction services in these areas, characterise the population in need and inform the immediate scale up of HIV prevention and treatment services.[14]

Clients who had migrated had 44% lower incidence of HIV compared to those who had not migrated. This is contrary to other studies that note higher HIV incidence among migrants particularly in the first two years of migration. [29] Notably, fewer people reported migration in in higher incidence border areas (5.8% vs 14.5%). One consequence of migration in the Myanmar context could be relocation to areas of increased security, including for health. Over half (52%) of data on migration was missing, we are therefore likely to have underestimated any association between migration and incidence. Our measure of migration was historic, and we are not able to examine whether incidence was higher among those who had more recently migrated. It was also broad and could not differentiate between forced displacement or migration for economic reasons. Findings from a survey of AHRN clients examining the effect of migration on symptoms of anxiety or depression suggested high rates of migration among clients, with 28.3% not living in the town their whole life, and high levels of past migration for economic reasons (77.9%) or as a result of war or armed conflict (19.5%).[30] Further research should consider the factors leading people to migrate, the different experience of vulnerability for those who migrate compared to those who do not.

The incidence of HIV among PWUD is lower than a recent study among men who have sex with men (MSM) and transgender populations suggests (10.1 per 100 person years). This was conducted among a small (n=279) sample in Yangon and Mandalay in Myanmar.[31] Our finding of higher incidence among those younger than 25 years (8.9/ 100 pyrs) is comparable with the MSM sample who were predominantly young (77% aged 25 years or younger compared to 5.2% in our programme sample). We found higher incidence among those who inject (6.8/100 pyrs) compared to those using drugs through non-injecting routes (1.2/100 pyrs). While this indicates that the epidemic is driven through injecting risk practices primarily, incidence is also high among people who use drugs via non-injecting routes. This points to the need to prioritise sexual risk reduction interventions at harm reduction programmes. Our estimates of incidence are in line with analyses of programmatic data from Médecins du Monde (MDM) providing harm reduction services in Kachin State that estimated incidence to be 7.1/100 pyrs (n=2277).[10] We found no evidence that incidence among clients declined over time in contrast to evidence from prospective cohorts of PWID in Thailand and the MDM programmatic data.[10, 32] Our analyses did not account for the provision of OAT or needle/syringes to PWID although both interventions are a cornerstone of AHRN’s HIV prevention activities and both OAT and needle/syringe provision are associated with reduced HIV incidence in Kachin and globally.[9, 10] At the point of analysis HIV testing data were not systematically linked to OAT uptake. Coverage of OAT in Myanmar is currently estimated to be 17% of the PWID population nationally, far lower than the WHO’s recommended guidelines of 40%.[33, 34] Other evidence suggests 1 in 5 PWID do not have sufficient clean needle/syringes for each injection.[35] There is emerging data from other borderland areas in the region of the effectiveness of cross-border interventions that involve implementation of dual interventions of needle/syringes and peer education in reducing HIV incidence among PWID in China and Vietnam.[25] High incidence and insufficient coverage of these key prevention interventions clearly point to the need to expand uptake and coverage of NSPs and OAT alongside use of pre-exposure prophylaxis and access to HIV treatment among PWID and ensuring interventions are delivered in borderland areas and between sites of frequent movement.

The strengths of our analysis lie in the large longitudinal sample of PWUD across a wide geographic area. Our findings provide the first estimate of HIV incidence in Myanmar among PWUD that focusses on geographic context and migration building on the growing evidence of the utility of programme data to estimate HIV incidence and illustrating their use in measuring structural factors in transmission.[23, 36] A key limitation is the measurement of exposure variables only at first registration, that does not necessarily reflect exposure at seroconversion. In relation to borderland and mining areas, individuals may have moved on by the time they engage in a second HIV test or no longer define themselves as migrants. Our measure of mining reflects risk in the wider community proximal to mines, rather than occupational hazards of working in mines or related activities. Only a third of people located in mining areas reported mining as their occupation. Industrial mines have been identified as potential hotspots for HIV transmission triggering changes in practices of local communities more conducive to HIV transmission due to migration of people for short term work from areas of high HIV prevalence, and greater propensity for people to engage with sex work or other risky sexual practices.[37] While our findings don’t indicate that the presence of mining activity substantially alters HIV vulnerability in the wider community of PWUD, we do observe a greater proportion of women and migrants in mining areas suggestive of changes to the community. Appropriate HIV responses need to be tailored to address the needs of these populations.

Analyses draw on a convenience sample from AHRN project sites and may not represent the overall population of PWUD, particularly given over a third were excluded due to not having an HIV test. Our sample is similar to othere including a community recruited sample of PWID conducted in 2017 across multiple sites who were also predominantly male (95.6% vs 98.2%) and used heroin (93.9% vs 98.8%).[13] Although our sample was older (median 36 vs 30 years) than the IBBS sample and other surveys [13, 38, 39] and with lower reported illiteracy (4.6% vs 15.7%).[39] National AIDS Programme guidelines recommend HIV testing every 6 months, but the median time between tests was 1.1 years among our sample with 38.3% of clients not being tested at all and only 26.4% with repeat tests. While this is low, it is in line with community surveys that suggest 51.6% of PWID had never had an HIV test and 48% were tested over one year ago.[13] Gender was measured as a binary, so our analyses fail to document additional risk among transgender populations. Only 4% of the sample were female. Further work is needed to engage women who use drugs in these services and document prevention and treatment needs.[7, 13] Our reliance on routine programmatic data with a limited number of indicators limits our understanding of the role of other structural factors or mediators, such as ethnicity for example that might contribute to conflict in borderland areas and HIV transmission. Missing data and lack of linkage to measures of OAT and ART uptake means it was not possible to control for all potential confounders in the analysis.

## Conclusions

Our findings contribute to the body of evidence that document the importance of borderland areas in disease transmission and the imperative to intensify harm reduction interventions with a focus on cross-border interventions. The recent introduction of PrEP among PWID in Myanmar is timely, but current pilot studies in harm reduction services are focussed on people who have injected in the last 6 months only, excluding people on OAT or using drugs via other routes. Our findings highlight the importance of extending eligibility criteria to include PWUD with multiple partners or engaging in unprotected sex in areas of high HIV incidence or prevalence in line with national guidelines for other key populations. Increasing uptake of HIV testing is imperative alongside the scale up of evidenced based interventions to address sexual and injecting risk practices including PrEP, distribution of condoms alongside needles/syringe distribution and OAT to curb the high rates of HIV transmission among these populations particularly among young people.

## Data Availability

Data cannot be shared publicly because the dataset contains individual level information on a population engaging in illegal activities (drug use) in a country currently led by a military dictatorship. The dataset contains data on demographic characteristics and geographical location of individuals that taken together could lead to deductive disclosure of individuals. If required a restricted dataset could be make available on request to AHRN the data holders following the implementation of a data sharing agreement and ethics approvals.

## Acknowledgement

We thank AHRN clients and staff for participation in the study and thanks to Sophia Garkov for assistance with the production of the map (figure 1).

## References

1. Publication UN. World Drug Report 2021. 2021.https://www.unodc.org/res/wdr2021/field/WDR21_Booklet_1.pdf

2. Degenhardt L, Peacock A, Colledge S, Leung J, Grebely J, Vickerman P, Stone J, Cunningham EB, Trickey A, Dumchev K, Lynskey M, Griffiths P, Mattick RP, Hickman M, Larney S. Global prevalence of injecting drug use and sociodemographic characteristics and prevalence of HIV, HBV, and HCV in people who inject drugs: a multistage systematic review. Lancet Glob Health. 2017;5(12):e1192–e207.10.1016/S2214-109X(17)30375-3

3. Mathers BM, Degenhardt L, Phillips B, Wiessing L, Hickman M, Strathdee SA, Wodak A, Panda S, Tyndall M, Toufik A, Mattick RP. Global epidemiology of injecting drug use and HIV among people who inject drugs: a systematic review. Lancet. 2008;372(9651):1733-45.10.1016/s0140-6736(08)61311-2

4. Degenhardt L, Mathers B, Guarinieri M, Panda S, Phillips B, Strathdee SA, Tyndall M, Wiessing L, Wodak A, Howard J. Meth/amphetamine use and associated HIV: Implications for global policy and public health. Int J Drug Policy. 2010;21(5):347–58.10.1016/j.drugpo.2009.11.007

5. United Nations Office on Drugs and Crime. World Drug Report 2016. Vienna; 2016.

6. Beyrer C, Razak MH, Lisam K, Chen J, Lui W, Yu XF. Overland heroin trafficking routes and HIV-1 spread in south and south-east Asia. AIDS. 2000;14(1):75–83. 10.1097/00002030-200001070-00009

7. O’Brien S, Kyaw KWY, Jaramillo MM, Roberts B, Bijl M, Platt L. Correction to: Determinants of health among people who use illicit drugs in the conflict-affected countries of Afghanistan, Colombia and Myanmar: a systematic review of epidemiological evidence. Conflict and Health. 2023;17(1):8.10.1186/s13031-023-00506-z

8. UNAIDS. Dangerous Inequalities. World AIDS Day report 2022. Geneva: Joint United Nations Programme on HIV/AIDS;; 2022.https://www.unaids.org/sites/default/files/media_asset/dangerous-inequalities_en.pdf

9. MacArthur GJ, Minozzi S, Martin N, Vickerman P, Deren S, Bruneau J, Degenhardt L, Hickman M. Opiate substitution treatment and HIV transmission in people who inject drugs: systematic review and meta-analysis. BMJ: British Medical Journal. 2012;345:e5945.10.1136/bmj.e5945

10. McNaughton AL, Stone J, Oo KT, Let ZZ, Taw M, Aung MT, Min AM, Lim AG, Wisse E, Vickerman P. Trends in HIV incidence following scale-up of harm reduction interventions among people who inject drugs in Kachin, Myanmar, 2008–2020: analysis of a retrospective cohort dataset. The Lancet Regional Health – Western Pacific.10.1016/j.lanwpc.2023.100718

11. Strathdee SA, Stockman JK. Epidemiology of HIV among injecting and non-injecting drug users: current trends and implications for interventions. Curr HIV/AIDS Rep. 2010;7(2):99–106. 10.1007/s11904-010-0043-7

12. Michel L, Nguyen LT, Nguyen AK, Ekwaru JP, Laureillard D, Nagot N, Phan O, Khuat OTH. Exposure to HIV risks among young people who use drugs (YPUD) in three cities in Vietnam: time to develop targeted interventions. Harm Reduction Journal. 2020;17(1):13. 10.1186/s12954-020-00357-4

13. National AIDS Program. Myanmar Integrated Biological and Behavioural Surveillance Survey & population size estimates people who inject drugs (PWID) 2017-2018. Ministry of Health and Sports, Myanmar; 2019 2019/01//.

14. Mining, drugs and conflict are stretching the AIDS response in northern Myanmar [press release]. UNAIDS 2020.

15. Williams CT, Liu W, Levy JA. Crossing over: drug network characteristics and injection risk along the China-Myanmar border. AIDS Behav. 2011;15(5):1011–6.10.1007/s10461-010-9764-2

16. Saw YM, Saw TN, Chan N, Cho SM, Jimba M. Gender-specific differences in high-risk sexual behaviors among methamphetamine users in Myanmar-China border city, Muse, Myanmar: who is at risk? BMC Public Health. 2018;18:209.10.1186/s12889-018-5113-6

17. Marmot M. Social determinants of health inequalities. Lancet. 2005;365(9464):1099-104.10.1016/s0140-6736(05)71146-6

18. Rhodes T. Risk environments and drug harms: a social science for harm reduction approach. Int J Drug Policy. 2009;20(3):193–201.10.1016/j.drugpo.2008.10.003

19. UN Office for the Coordination of Humanitarian Affairs. Myanmar Humanitarian Update No 17. UN Office for the Coordination of Humanitarian Affairs; 2022.https://reliefweb.int/report/myanmar/myanmar-humanitarian-update-no-17-19-april-2022

20. Wang R-B, Dong J-Q, Xia Z-G, Cai T, Zhang Q-F, Zhang Y, Tian Y-H, Sun X-Y, Zhang G-Y, Li Q-P, Xu X-Y, Li J-Y, Zhang J. Lessons on malaria control in the ethnic minority regions in Northern Myanmar along the China border, 2007–2014. Infectious Diseases of Poverty. 2016;5(1):95.10.1186/s40249-016-0191-0

21. Zhou Y-H, Liu F-L, Yao Z-H, Duo L, Li H, Sun Y, Zheng Y-T. Comparison of HIV-, HBV-, HCV- and Co-Infection Prevalence between Chinese and Burmese Intravenous Drug Users of the China-Myanmar Border Region. PLOS ONE. 2011;6(1):e16349.10.1371/journal.pone.0016349

22. Unit MIM. MIMU Township Profiles Yangon, Myanmar 2023 [Available from: https://themimu.info/.

23. Hargreaves JR, Mtetwa S, Davey C, Dirawo J, Chidiya S, Benedikt C, Naperiela Mavedzenge S, Wong-Gruenwald R, Hanisch D, Magure T, Mugurungi O, Cowan FM. Implementation and Operational Research: Cohort Analysis of Program Data to Estimate HIV Incidence and Uptake of HIV-Related Services Among Female Sex Workers in Zimbabwe, 2009-2014. J Acquir Immune Defic Syndr. 2016;72(1):e1-8.10.1097/qai.0000000000000920

24. Li L, Assanangkornchai S, Duo L, McNeil E, Li J. Cross-border activities and association with current methamphetamine use among Chinese injection drug users (IDUs) in a China– Myanmar border region. Drug and Alcohol Dependence. 2014;138:48–53.10.1016/j.drugalcdep.2014.01.021

25. Hammett TM, Des Jarlais DC, Kling R, Kieu BT, McNicholl JM, Wasinrapee P, McDougal JS, Liu W, Chen Y, Meng D, Doan N, Tho HN, Quyen NH, Tren VH. Controlling HIV epidemics among injection drug users: eight years of Cross-Border HIV prevention interventions in Vietnam and China. PLOS ONE. 2012;7(8):e43141.10.1371/journal.pone.0043141

26. Hammett TM, Johnston P, Kling R, Liu W, Ngu D, Tung ND, Binh KT, Dong HV, Hoang TV, Van LK, Donghua M, Chen Y, Des Jarlais DC. Correlates of HIV status among injection drug users in a border region of southern China and northern Vietnam. J Acquir Immune Defic Syndr. 2005;38(2):228–35.10.1097/00126334-200502010-00016

27. Pollini RA, Strathdee SA. Indicators of methamphetamine use and abuse in San Diego County, California: 2001-2005. J Psychoactive Drugs. 2007;Suppl 4:319-25.10.1080/02791072.2007.10399893

28. Aung YK, Zin SS, Tesfazghi K, Paudel M, Thet MM, Thein ST. A comparison of malaria prevention behaviours, care-seeking practices and barriers between malaria at-risk worksite migrant workers and villagers in Northern Shan State, Myanmar—a mixed method study. Malaria Journal. 2022;21(1):162.10.1186/s12936-022-04193-8

29. Olawore O, Tobian AAR, Kagaayi J, Bazaale JM, Nantume B, Kigozi G, Nankinga J, Nalugoda F, Nakigozi G, Kigozi G, Gray RH, Wawer MJ, Ssekubugu R, Santelli JS, Reynolds SJ, Chang LW, Serwadda D, Grabowski MK. Migration and risk of HIV acquisition in Rakai, Uganda: a population-based cohort study. The Lancet HIV. 2018;5(4):e181-e9.10.1016/S2352-3018(18)30009-2

30. Kyaw KWY, Platt L, Bijl M, Rathod SD, Naing AY, Roberts B. The effect of different types of migration on symptoms of anxiety or depression and experience of violence among people who use or inject drugs in Kachin State, Myanmar. Harm Reduction Journal. 2023;20(1):45.10.1186/s12954-023-00766-1

31. Veronese V, Traeger M, Oo ZM, Tun TT, Oo NN, Maung H, Hughes C, Pedrana A, Stoové M. HIV incidence and factors associated with testing positive for HIV among men who have sex with men and transgender women in Myanmar: data from community-based HIV testing services. Journal of the International AIDS Society. 2020;23(2):e25454.10.1002/jia2.25454

32. Martin M, Vanichseni S, Sangkum U, Mock PA, Leethochawalit M, Chiamwongpaet S, Pitisuttithum P, Kaewkungwal J, van Griensven F, McNicholl JM, Tappero JW, Mastro TD, Kittimunkong S, Choopanya K. HIV Incidence and Risk Behaviours of People Who Inject Drugs in Bangkok, 1995–2012. eClinicalMedicine. 2019;9:44-51.10.1016/j.eclinm.2019.03.012

33. World Health Organisation, UNODC, UNAIDS. Technical Guide for Countries to Set Targets for Universal Access to HIV Prevention, Treatment and Care for Injecting Drug Users. Geneva, Switzerland: World Health Organisation 2012.https://www.who.int/publications/i/item/978924150437

34. Ministry of Health and Sports. Progress Report 2018. National AIDS Program. Myanmar: Ministry of Health and Sports; 2018.https://www.thelancet.com/action/showPdf?pii=S2666-6065%2823%2900036-6

35. O’Keefe D, Aung SM, Pasricha N, Wun T, Linn SK, Lin N, Aitken C, Hughes C, Dietze P. Measuring individual-level needle and syringe coverage among people who inject drugs in Myanmar. International Journal of Drug Policy. 2018;58:22–30.10.1016/j.drugpo.2018.04.010

36. World Health Organisation. Consolidated guidelines on person-centred HIV strategic information: strengthening routine data for impact. Geneva: World Health Organisation; 2022.

37. Dietler D, Farnham A, Lyatuu I, Fink G, Winkler MS. Industrial mining and HIV risk: evidence from 39 mine openings across 16 countries in sub-Saharan Africa. AIDS. 2022;36(11):1573–81.10.1097/QAD.0000000000003294

38. Swe LA, Rashid A. HIV prevalence among the female sex workers in major cities in Myanmar and the risk behaviors associated with it. HIV AIDS (Auckl). 2013;5:223–30.10.2147/HIV.S50171

39. Swe LA, Nyo KK, Rashid AK. Risk behaviours among HIV positive injecting drug users in Myanmar: a case control study. Harm Reduction Journal. 2010;7(12).10.1186/1477-7517-7-12

